# Risk factors for illness severity among pregnant women with confirmed SARS-CoV-2 infection – Surveillance for Emerging Threats to Mothers and Babies Network, 20 state, local, and territorial health departments, March 29, 2020 -January 8, 2021

**DOI:** 10.1101/2021.02.27.21252169

**Authors:** Romeo R. Galang, Suzanne M. Newton, Kate R. Woodworth, Isabel Griffin, Titilope Oduyebo, Christina L. Sancken, Emily O’Malley Olsen, Kathy Aveni, Heather Wingate, Hanna Shephard, Chris Fussman, Zahra S. Alaali, Samantha Siebman, Umme-Aiman Halai, Camille Delgado Lopez, Jerusha Barton, Mamie Lush, Paul H. Patrick, Levi Schlosser, Ayomide Sokale, Ifrah Chaudhary, Bethany Reynolds, Similoluwa Sowunmi, Nicole Gaarenstroom, Jennifer S. Read, Sarah Chicchelly, Leah de Wilde, Eduardo Azziz-Baumgartner, Aron J. Hall, Van T. Tong, Sascha Ellington, Suzanne M. Gilboa, CDC COVID-19 Response Pregnancy and Infant Linked Outcomes Team

## Abstract

**Background:** Pregnant women with coronavirus disease 2019 (COVID-19) are at increased risk for severe illness compared with nonpregnant women. Data to assess risk factors for illness severity among pregnant women with COVID-19 are limited. This study aimed to determine risk factors associated with COVID-19 illness severity among pregnant women with SARS-CoV-2 infection.

**Methods:** Pregnant women with SARS-CoV-2 infection confirmed by molecular testing were reported during March 29, 2020–January 8, 2021 through the Surveillance for Emerging Threats to Mothers and Babies Network (SET-NET). Criteria for illness severity (asymptomatic, mild, moderate-to-severe, or critical) were adapted from National Institutes of Health and World Health Organization criteria. Crude and adjusted risk ratios for moderate-to-severe or critical COVID-19 illness were calculated for selected demographic and clinical characteristics.

**Results:** Among 5,963 pregnant women with SARS-CoV-2 infection, moderate-to-severe or critical COVID-19 illness was associated with age 30–39 years, Black/Non-Hispanic race/ethnicity, healthcare occupation, pre-pregnancy obesity, chronic lung disease, chronic hypertension, cardiovascular disease, and pregestational diabetes mellitus. Risk of moderate-to-severe or critical illness increased with the number of underlying medical or pregnancy-related conditions.

**Conclusions:** Pregnant women with moderate-to-severe or critical COVID-19 illness were more likely to be older and have underlying medical conditions compared to pregnant women with asymptomatic infection or mild COVID-19 illness. This information might help pregnant women understand their risk for moderate-to-severe or critical COVID-19 illness and inform targeted public health messaging.

**Summary:** Among pregnant women with COVID-19, older age and underlying medical conditions were risk factors for increased illness severity. These findings can be used to inform pregnant women about their risk for severe COVID-19 illness and public health messaging.

## Introduction

Pregnant women with coronavirus disease 2019 (COVID-19) are at increased risk for severe illness compared with nonpregnant women [1]. A limited number of studies have suggested that risk factors for severe COVID-19 illness, such as older age and underlying medical conditions, might be similar between pregnant and non-pregnant people; however, individual studies have been limited in sample size, varied in sampling frame and inclusion criteria (e.g., inclusion of women with suspected COVID-19 and/or those with confirmed COVID-19), and primarily reported on pregnant women requiring hospitalization (including for childbirth) [2-4]. Additional information on risk factors for severe COVID-19 illness are needed to inform discussions about risk for severe illness, to guide public health messaging and to inform decision-making around resource allocation.

Public health jurisdictions report information, including pregnancy status, on confirmed and probable COVID-19 cases to CDC through the National Notifiable Diseases Surveillance System [5]. Through the Surveillance for Emerging Threats to Mothers and Babies Network (SET-NET), health departments from 20 jurisdictions collected supplementary information on pregnancy outcomes among women with SARS-CoV-2 infection confirmed by nucleic acid amplification testing and reported during March 29, 2020–January 8, 2021 [6]. To determine risk factors associated with COVID-19 illness severity, demographic and selected clinical characteristics were compared between pregnant women with moderate-to-severe or critical illness and those with asymptomatic infection or mild illness.

## Materials and Methods

SET-NET is longitudinal surveillance of pregnant women and their infants to understand the effects of emerging and reemerging threats [6]. Supplementary pregnancy-related information is reported for women with laboratory confirmed SARS-CoV-2 infection (based on detection of SARS-CoV-2 in a clinical specimen by nucleic acid amplification testing) during pregnancy through the day of delivery in 2020 [7]. As of January 8, 2021, health departments from 20 jurisdictions (California [excluding Los Angeles County], Georgia, Houston, Kansas, Los Angeles County, Massachusetts, Michigan, Minnesota, Nebraska, Nevada, New Jersey, New York [excluding New York City], North Dakota, Oklahoma, Pennsylvania [excluding Philadelphia], Philadelphia, Puerto Rico, Tennessee, U.S. Virgin Islands, and Vermont) have contributed data [6]. Pregnancy status was ascertained through routine COVID-19 case surveillance or through matching of reported cases with other data sources (e.g., vital records, administrative data) to identify unreported pregnancy status or verify pregnancy status. Data were abstracted using standard data elements; sources include routine public health investigations, vital records, laboratory reports, and medical records. SET-NET methodology has been previously described [6].

Criteria for illness severity (asymptomatic, mild, moderate-to-severe, or critical) were adapted from National Institutes of Health and World Health Organization severity of illness categories (Figure) [8-9]. Women were considered asymptomatic if reported as having an absence of symptoms using a symptom status variable. Criteria were applied to classify severity using submitted data (including symptoms, intensive care unit (ICU) admission, invasive ventilation, use of COVID-19 therapies, complications associated with COVID-19, and death). If data were not reported for an outcome, the outcome was assumed not to have occurred. Crude risk ratios (RR) for moderate-to-severe or critical illness were calculated for selected demographic characteristics within age group, race/ethnicity, health insurance type, healthcare worker status and selected clinical characteristics, including diagnosis of underlying medical condition (pre-pregnancy obesity [body mass index ≥30 kg/m], chronic lung disease, chronic hypertension, pregestational diabetes mellitus, cardiovascular disease, and immunosuppression), trimester of SARS-CoV-2 infection, and diagnosis of pregnancy-related condition (gestational diabetes and gestational hypertension) as reported through contact tracing, vital statistics, or medical records, compared to selected referent groups [6]. We also calculated crude risk ratios comparing risk of moderate-to-severe or critical illness among pregnant women with any one condition (underlying medical or pregnancy-related condition), any two conditions, and three or more conditions compared to those without report of any condition. Adjusted risk ratios (aRR) and 95% CIs for moderate-to-severe or critical illness were estimated by binomial regression with the log link function, accounting for age (in years) as a continuous variable. Analyses were conducted using SAS (version 9.4; SAS Institute). This activity was reviewed by CDC, determined to be a non-research, public health surveillance activity, and was conducted consistent with applicable federal law and CDC policy [10].

**Figure.**
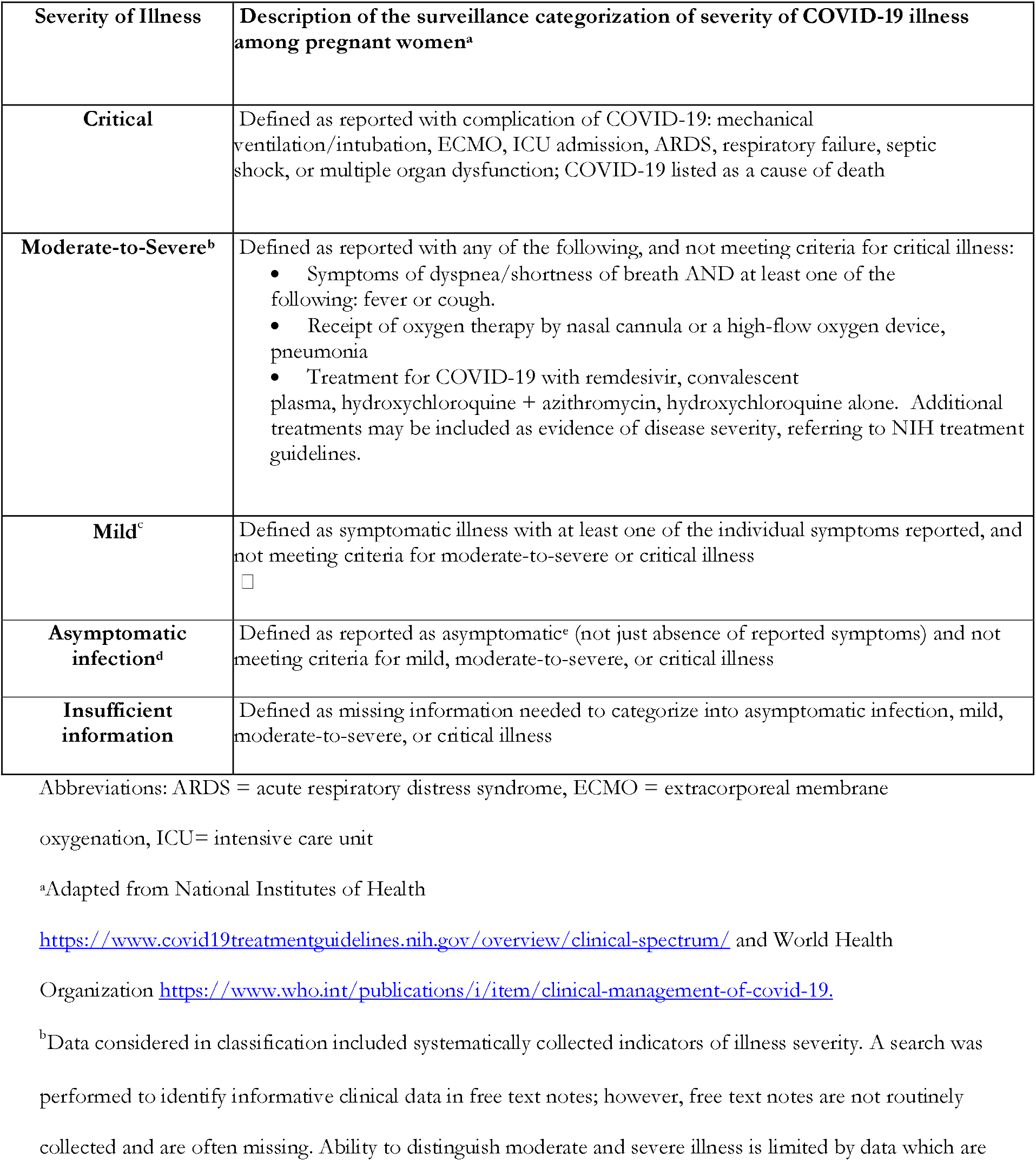
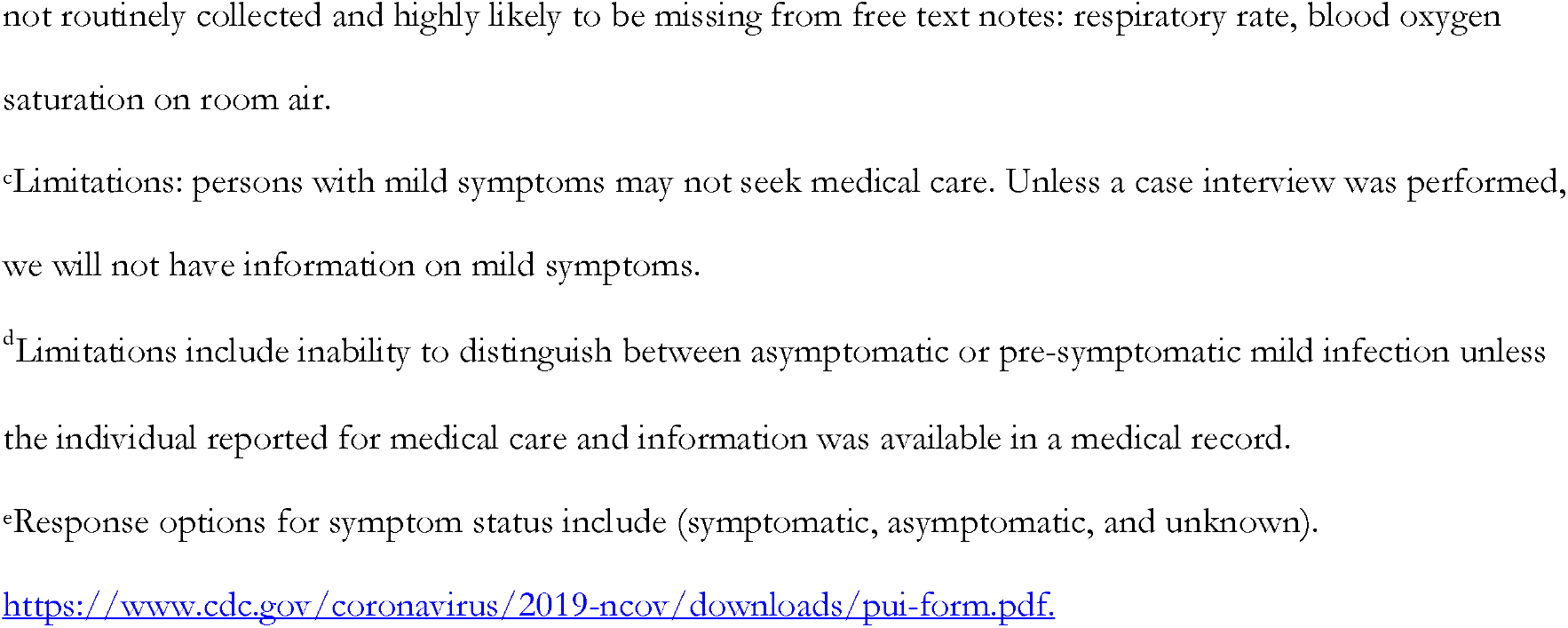
Criteria for categorizing severity of illness among pregnant women with SARS-CoV-2 infection, Surveillance for Emerging Threats to Mothers and Babies Network (SET-NET)

## Results

During March 29–January 8, 2021, data for 10,996 pregnant women with confirmed SARS-CoV-2 infection were submitted to SET-NET. Data for 5,033 (45.8%) women were insufficient for categorizing illness severity. The remainder of this report focuses on 5,963 (54.2%) pregnant women with SARS-CoV-2 infection and sufficient information to categorize illness severity.

Most women were aged 20-39 years (90.8%), 44.9% were Hispanic or Latina (Hispanic) ethnicity, and 55.7% had Medicaid (Table 1). At least one underlying medical condition was reported for 1,827 (35.3%) women, with pre-pregnancy obesity (27.2%) most commonly reported. Gestational diabetes was reported in 8.9% of women and gestational hypertension in 9.0%. Most women had SARS-CoV-2 infection identified in the third (60.6%) or second (29.1%) trimester (based on date of first positive test or symptom onset).

**Table 1.** Risk ratios for moderate-to-severe or critical illness among pregnant women with SARS-CoV-2 infection during pregnancy compared to asymptomatic infection or mild illness for selected demographic and clinical characteristics, Surveillance for Emerging Threats to Mothers and Babies Network, 20 state, local, and territorial health departments, March 29, 2020 -January 8, 2021 (n=5,963)^a^

|  | No. of women (%) |  |  | Risk Ratio | 95% CI | Adjusted Risk Ratio | 95% CI |
| --- | --- | --- | --- | --- | --- | --- | --- |
|  | [Total no. of women with available information] |  |  |  |  |  |  |
|  | Total | Moderate-to-severe or critical illness | Asymptomatic infection or mild illness |  |  |  |  |
| **Total** | 5,963 | 1,379 (23.1) | 4,584 (76.9) |  |  |  |  |
| **Age** | [4215] | [1032] | [3183] |  |  |  |  |
| <20 | 249 (5.9) | 49 (4.8) | 200 (6.3) | 1 | ref. | - | - |
| 20-24 | 847 (20.1) | 182 (17.6) | 665 (20.9) | 1.09 | (0.82, 1.44) | - | - |
| 25-29 | 1306 (31.0) | 307 (29.8) | 999 (31.4) | 1.19 | (0.91, 1.56) | - | - |
| 30-34 | 1098 (26.1) | 292 (28.3) | 806 (25.3) | **1.35** | **(1.03, 1.77)** | - | - |
| 35-39 | 577 (13.7) | 163 (15.8) | 414 (13.0) | **1.44** | **(1.08, 1.90)** | - | - |
| 40 + | 138 (3.3) | 39 (3.8) | 99 (3.1) | 1.44 | (1.00, 2.07) | - | - |
| **Race/ethnicity** | [4961] | [1171] | [3790] |  |  |  |  |
| White, Non-Hispanic | 1494 (30.1) | 340 (29.0) | 1154 (30.5) | 1 | ref. | 1 | ref. |
| Asian, Non-Hispanic | 178 (3.6) | 50 (4.3) | 128 (3.4) | 1.23 | (0.96, 1.59) | 1.27 | (0.95, 1.68) |
| Black, Non-Hispanic | 898 (18.1) | 227 (19.4) | 671 (17.7) | 1.11 | (0.96, 1.29) | **1.22** | **(1.04, 1.43)** |
| Hispanic or Latina | 2225 (44.9) | 526 (44.9) | 1699 (44.8) | 1.04 | (0.92, 1.17) | 0.92 | (0.80, 1.05) |
| Multiple or other, Non-Hispanic | 166 (3.4) | 28 (2.4) | 138 (3.6) | 0.74 | (0.52, 1.05) | 1.02 | (0.71, 1.46) |
| **Health insurance** | [3735] | [931] | [2804] |  |  |  |  |
| Private | 1451 (38.9) | 342 (36.7) | 1109 (39.6) | 1 | ref. | 1 | ref. |
| Medicaid | 2079 (55.7) | 545 (58.5) | 1534 (54.7) | 1.11 | (0.99, 1.25) | 1.13 | (0.99, 1.29) |
| Other | 130 (3.5) | 26 (2.8) | 104 (3.7) | 0.85 | (0.59, 1.21) | 1.07 | (0.75, 1.51) |
| Self-pay/none | 75 (2.0) | 18 (1.9) | 57 (2.0) | 1.02 | (0.67, 1.54) | 0.95 | (0.61, 1.47) |
| **Healthcare occupation** | [3245] | [806] | [2439] |  |  |  |  |
| No | 2512 (77.4) | 579 (71.8) | 1933 (79.3) | 1 | ref. | 1 | ref. |
| Yes | 733 (22.6) | 227 (28.2) | 506 (20.8) | **1.34** | **(1.18, 1.53)** | **1.31** | **(1.14, 1.50)** |
| **Trimester of SARS-CoV-2 infection<sup>c</sup>** | [5100] | [1216] | [3884] |  |  |  |  |
| First | 525 (10.3) | 123 (10.1) | 402 (10.4) | 1 | ref. | 1 | ref. |
| Second | 1486 (29.1) | 379 (31.2) | 1107 (28.5) | 1.09 | (0.91, 1.30) | 1.01 | (0.82, 1.24) |
| Third | 3089 (60.6) | 714 (58.7) | 2375 (61.2) | 1 | (0.83, 1.17) | 1.02 | (0.84, 1.24) |
| **Underlying medical condition<sup>d</sup>** | [5177] | [1144] | [4033] |  |  |  |  |
| Any underlying medical condition | 1827 (35.3) | 498 (43.5) | 1329 (33.0) | **1.41** | **(1.28, 1.57)** | **1.48** | **(1.32, 1.67)** |
| Obesity<sup>e</sup> | 1407 (27.2) | 373 (32.6) | 1034 (25.6) | **1.32** | **(1.17, 1.48)** | **1.29** | **(1.12, 1.48)** |
| Chronic lung disease | 318 (6.1) | 102 (8.9) | 216 (5.4) | **1.39** | **(1.16, 1.66)** | **1.44** | **(1.19, 1.73)** |
| Chronic hypertension | 164 (3.2) | 51 (4.5) | 113 (2.8) | **1.37** | **(1.09, 1.74)** | **1.35** | **(1.05, 1.73)** |
| Diabetes mellitus (type 1 or type 2) | 117 (2.3) | 41 (3.6) | 76 (1.9) | **1.54** | **(1.20, 1.98)** | **1.57** | **(1.21, 2.04)** |
| Cardiovascular disease | 86 (1.7) | 30 (2.6) | 56 (1.4) | **1.54** | **(1.14, 2.08)** | **1.46** | **(1.06, 2.02)** |
| Immunosuppression | 41 (0.8) | 11 (1.0) | 30 (0.7) | 1.28 | (0.76, 2.12) | 1.49 | (0.88, 2.53) |
| **Pregnancy-related condition<sup>d</sup>** | [4470] | [995] | [3475] |  |  |  |  |
| Gestational diabetes | 447 (10.0) | 119 (12.0) | 328 (9.4) | **1.23** | **(1.04, 1.45)** | 1.1 | (0.90, 1.33) |
| Gestational hypertension | 452 (10.1) | 115 (11.6) | 337 (9.7) | 1.16 | (0.98, 1.38) | 1.11 | (0.92, 1.35) |
| **Number of conditions<sup>f</sup>** | [5337] | [1166] | [4171] |  |  |  |  |
| None | 3106 (58.2) | 589 (50.5) | 2517 (60.4) | 1 | ref. | 1 | ref. |
| Any 1 condition | 1602 (30.0) | 381 (32.7) | 1221 (29.3) | **1.25** | **(1.12, 1.40)** | **1.3** | **(1.13, 1.48)** |
| Any 2 conditions | 505 (9.5) | 146 (12.5) | 359 (8.6) | **1.52** | **(1.31, 1.78)** | **1.59** | **(1.33, 1.89)** |
| Any 3 or more conditions | 124 (2.3) | 50 (4.3) | 74 (1.8) | **2.13** | **(1.70, 2.67)** | **1.82** | **(1.39, 2.39)** |

<sup>a</sup>During March 29, 2020–January 8, 2021, data for 10,996 pregnant women with SARS-CoV-2 infection were submitted to SET-NET. Data for 5,033
(45.8%) women were insufficient for categorizing severity of illness.
<sup>b</sup>Adjusted for age as a continuous variable.
<sup>c</sup>Trimester of SARS-CoV-2 infection based on date of first positive test or symptom onset.
<sup>d</sup>Referent group: pregnant women without report of underlying medical condition or pregnancy-related condition, respectively.
<sup>e</sup>Pregestational obesity defined as body mass index $\geq 30$ kg/m<sup>2</sup>.
<sup>f</sup>Includes underlying medical conditions (pre-pregnancy obesity, chronic lung disease, chronic hypertension, diabetes mellitus (type 1 or type 2),
cardiovascular disease and immunosuppression) and pregnancy-related conditions (gestational diabetes and gestational hypertension

In crude analysis, pregnant women who were 30-34 years (RR=1.35, 95% CI: 1.03, 1.77) and 35-39 years of age (RR=1.44, 95% CI: 1.08, 1.90) were at an increased risk of moderate-to-severe or critical illness compared to pregnant women who were <20 years of age. Pregnant women reported as having a healthcare occupation (RR=1.34, 95% CI: 1.18, 1.53) were at an increased risk of moderate-to-severe or critical illness compared to pregnant women who were not reported as being in a healthcare occupation. Pregnant women with pre-pregnancy obesity (RR=1.32, 95% CI: 1.17, 1.48), chronic lung disease (RR=1.39, 95% CI: 1.16, 1.66), chronic hypertension (RR=1.37, 95% CI: 1.09, 1.74), cardiovascular disease (RR=1.54, 95% CI: (1.14, 2.08), pregestational diabetes mellitus (RR=1.54, 95% CI: 1.20, 1.98), and gestational diabetes (RR=1.23, 95% CI: 1.04, 1.45) were at increased risk of moderate-to-severe or critical illness compared to pregnant women without these conditions.

Presence of any health condition (underlying medical or pregnancy-related health condition) was associated with 25% increased risk (RR=1.25, 95% CI: 1.12, 1.40), two conditions was associated with a 52% increased risk (RR=1.52, 95% CI: 1.31, 1.78), and three or more conditions was associated with twice the risk (RR=2.13, 95% CI: 1.70, 2.67) of moderate-to-severe or critical illness compared to women without any reported conditions. Age younger than 29 years, age 30 to 39 years, race/ethnicity, health insurance type, trimester of SARS-CoV-2 infection, immunosuppression, and gestational hypertension were not associated with increased risk of moderate-to-severe or critical illness compared to the referent groups.

Adjusted risk ratios were similar to crude risk ratios with two exceptions. After adjustment for age as a continuous variable, Black/Non-Hispanic race/ethnicity was associated with a 22% increased risk (aRR=1.22, 95% CI: 1.04, 1.43) of moderate-to-severe or critical illness compared to the referent group (White, Non-Hispanic race/ethnicity), and gestational diabetes was not found to be associated with increased risk.

## Discussion

In an analysis of a large cohort of pregnant women with SARS-CoV-2 infection reported from health departments from 20 jurisdictions through SET-NET, age 30-39 years, Black/Non-Hispanic race/ethnicity, being a healthcare worker, and presence of any underlying medical condition were associated with increased risk of moderate-to-severe or critical illness. The number of underlying medical or pregnancy-related conditions demonstrated an exposure-response relation with risk for moderate-to-severe or critical illness. Data collection is ongoing, and findings may change as additional data are collected and analyzed. Data are reported by health departments and can be updated as new information becomes available. Enhanced efforts to improve reporting of clinical data related to illness severity are ongoing.

These findings of association between older age, Black/Non-Hispanic race/ethnicity, healthcare occupation, any underlying medical condition and increased risk of moderate-to-severe or critical COVID-19 illness are similar to those observed among nonpregnant adults. There have been few studies focused on risk factors for COVID-19 illness severity in pregnant women; those study findings suggest similar associations with older age and medical comorbidities as seen in the general adult population [2-4]. An association was not found with trimester of SARS-CoV-2 infection, similar to findings from a recent systematic review and meta-analysis of SARS-CoV-2 infection in pregnancy [4]. An association of Hispanic or Latina race/ethnicity with moderate-to-severe or critical illness was not identified; however, Hispanic or Latina women represented half of all women with moderate-to-severe or critical illness in this analysis.

The findings in this report are subject to at least four limitations. First, the clinical criteria for classifying illness severity in this analysis were adapted for surveillance purposes from existing frameworks and used severity indicators that were captured systematically, while other criteria may not have been captured (e.g., respiratory rate and oxygen saturation on room air). Misclassification of illness severity is possible, particularly when data to classify cases into moderate-to-severe or critical illness categories are missing, which might bias towards a lower severity classification and attenuate associations [11]. Similarly, data cannot distinguish between asymptomatic or pre-symptomatic mild infection unless the individual subsequently reported for medical care and information was available in a medical record. Additionally, women who were tested upon hospital admission for delivery may have developed more severe symptoms later on that were not captured by SET-NET. Among women with date of testing and outcome available, 26% were identified within two days of delivery, which could reflect universal screening on admission. Second, a large portion of women could not be categorized for illness severity due to insufficient information, and testing and reporting might be more frequent among women with more severe illness. The ability to detect differences in demographic characteristics between included and excluded women were limited by a large portion of missing demographic information among excluded cases due to the large surge of cases and limited capacity for complete data collection. Additionally, obtaining accurate data to distinguish underlying medical conditions from pregnancy-related medical conditions (e.g., diabetes vs gestational diabetes) depends on medical record abstraction. Potential misclassification of underlying medical conditions and pregnancy-related medical conditions might limit detection of an association with moderate-to-severe or critical illness. Medical record abstraction of clinical information are ongoing, and statistical comparisons by illness severity should be interpreted with caution. Third, while these data are population-based for the jurisdictions included, they are not nationally representative and include a higher frequency of Hispanic and Latina women compared with all women of reproductive age in national case surveillance data and with provisional national 2020 data on births among women with COVID-19 during pregnancy [1, 12-13]. Fourth, relative to the proportion of women with SARS-CoV-2 infection in the second and third trimesters of pregnancy, few women with first trimester infection have been reported to date. This limits our ability to understand whether trimester of infection is associated with severity of COVID-19 illness.

Despite these limitations, this report has several strengths, including the large size of the population-based cohort, inclusion of both hospitalized and non-hospitalized pregnant women, restriction of the study population to pregnant women with confirmed COVID-19, information to describe risk factors for illness severity among pregnant women with COVID-19, and uniform application of illness severity criteria.

Future research could further focus on clinical relevance of maternal COVID-19 illness severity and outcomes among newborns, infants, and children. Additional follow-up data on SARS-CoV-2 infection are needed to increase certainty of findings related to severity of COVID-19 illness and timing of infection during pregnancy.

These data can help counsel pregnant women about their risk for moderate-to-severe or critical COVID-19 illness and guide their choice of prevention strategies, target public health messaging, and inform decisions around resource allocation. It is important that pregnant women are informed of their increased risk for severe COVID-19 illness, the signs of severe COVID-19 illness, and strategies for prevention, including vaccination [14-16].

## Supporting information

STROBE Checklist

## Data Availability

These data are collected under relevant provisions of the Public Health Service Act and are protected at CDC by an Assurance of Confidentiality (Section 308(d) of the Public Health Service Act, 42 U.S.C. section 242 m(d)) (https://www.cdc.gov/od/science/integrity/confidentiality/), which prohibits use or disclosure of any identifiable or potentially identifiable information collected under the Assurance for purposes other than those set out in the Assurance. Publicly available aggregated data are available: https://www.cdc.gov/coronavirus/2019-ncov/cases-updates/special-populations/birth-data-on-covid-19.html. Requests for access will be considered on a case by case basis, and inquiries should be directed to

## Funding

This study was performed as regular work of the Centers for Disease Control and Prevention. This work is supported by the Epidemiology and Laboratory Capacity for Prevention and Control of Emerging Infectious Diseases (ELC) Cooperative Agreement (ELC CK19-1904) and through contractual mechanisms, including the Local Health Department Initiative.

## Acknowledgments

Joy Rende, Lindsey Sizemore, Elizabeth Harvey, Nicole D. Longcore, Nadia Thomas, Pauline Santos, Deirdre Depew, Jamie N. Sommer, Claire McGarry, Ona Loper, Shannon Baack, Miguel Valencia Prado, Leishla Nieves Ferrer, Mariam Marcano Huertas, Stephany Perez González, Glorimar Meléndez Rosario, Marangelí Olán Martínez, Hilcon Agosto Rosa, Reynaldo Pérez Alices, J. Michael Bryan, Cristina Meza, Victoria Sanon, Teri’ Willabus, Cynthia Carpentieri, Michael Andrews, Skip Frick, Robin M. Williams, Samir Koirala, Tyler Faulkner, Shannon Lawrence, Erika Fuchs, Celeste Illian, Elizabeth Wyckoff, Aasta D. Mehta, Patrick Nwachukwu, Lauren Orkis, Richard Olney, Valorie Eckert, Barbara Warmerdam, Olga Barer, State, local, and territorial health department personnel; U.S. clinical, public health, and emergency response staff members; CDC Epidemiology and Surveillance Task Force, CDC Data Analytics and Modeling Task Force

## COVID-19 Pregnancy and Infant Linked Outcomes Team (PILOT)

Jennifer Beauregard, CDC; Jason Hsia, CDC; Kellianne King, CDC; Jean Ko, CDC; Elizabeth Lewis, CDC; Susan Manning, CDC; Varsha Neelam, CDC; Mirna Perez, CDC; Emily Petersen, CDC; Megan Reynolds, CDC; Aspen Riser, CDC; Maria Rivera, CDC; Nicki Roth, CDC; Regina Simeone, CDC; John Sims, CDC; Ashley Smoots, CDC; Margaret Snead, CDC; Penelope Strid, CDC; Diana Valencia, CDC; Bailey Wallace, CDC; Tineka Yowe-Conley, CDC; Laura Zambrano, CDC; Lauren Zapata, CDC; Amanda Akosa, Eagle Global Scientific; John F. Nahabedian III, Eagle Global Scientific; Amitsingh Rathore, Eagle Global Scientific; Neha Shinde, Eagle Global Scientific; Veronica Burkel, Eagle Medical; Dena Cherry, General Dynamics Informational Technology; Daniel Chang, Oak Ridge Institute for Science and Education; Charise Fox, Oak Ridge Institute for Science and Education; Emily Reeves, Oak Ridge Institute for Science and Education; Ayzsa Tannis, Oak Ridge Institute for Science and Education; Susanna Trost, Oak Ridge Institute for Science and Education

## Potential conflict of interests

All authors have completed and submitted the International Committee of Medical Journal Editors form for disclosure of potential conflicts of interest. No potential conflicts of interest were disclosed. This work has not been published previously and is not under consideration for publication elsewhere.

## Notes

### Competing Interest Statement

The authors have declared no competing interest.

### Clinical Trial

This activity was reviewed by CDC, determined to be a non-research, public health surveillance activity, and was conducted consistent with applicable federal law and CDC policy.

### Clinical Protocols

https://www.ncbi.nlm.nih.gov/pmc/articles/PMC7643898/

### Author Declarations

This activity was reviewed by the human subjects advisor of the U.S. Centers for Disease Control and Prevention (CDC), National Center on Birth Defects and Developmental Disorders and was determined to be non-research, public health surveillance and exempt from IRB review. This activity was conducted consistent with applicable federal law and CDC policy. (Department of Health and Human Services - 45 C.F.R. part 46, 21 C.F.R. part 56; 42 U.S.C. Sect. 241(d); 5 U.S.C. Sect. 552a; 44 U.S.C. Sect. 3501 et seq. Available from: https://www.hhs.gov/ohrp/sites/default/files/ohrp/policy/ohrpregulations.pdf.)

