## Supplementary material for "Risk factors for illness severity among pregnant women with confirmed SARS-CoV-2 infection – Surveillance for Emerging Threats to Mothers and Babies Network, 20 state, local, and territorial health departments, March 29, 2020 -January 8, 2021": STROBE Checklist

### Reporting checklist for cohort study.

Based on the STROBE cohort guidelines.

#### Instructions to authors

Complete this checklist by entering the page numbers from your manuscript where readers will find each of the items listed below.

Your article may not currently address all the items on the checklist. Please modify your text to include the missing information. If you are certain that an item does not apply, please write "n/a" and provide a short explanation.

Upload your completed checklist as an extra file when you submit to a journal.

In your methods section, say that you used the STROBE cohortreporting guidelines, and cite them as:

von Elm E, Altman DG, Egger M, Pocock SJ, Gotzsche PC, Vandenbroucke JP. The Strengthening the Reporting of Observational Studies in Epidemiology (STROBE) Statement: guidelines for reporting observational studies.

|  |  | Reporting Item | Page Number |
| --- | --- | --- | --- |
| **Title and abstract** |  |  |  |
| Title | [#1a](https://www.goodreports.org/reporting-checklists/strobe-cohort/info/#1a) | Indicate the study’s design with a commonly used term in the title or the abstract | 1 |
| Abstract | [#1b](https://www.goodreports.org/reporting-checklists/strobe-cohort/info/#1b) | Provide in the abstract an informative and balanced summary of what was done and what was found | 4 |
| **Introduction** |  |  |  |
| Background / rationale | [#2](https://www.goodreports.org/reporting-checklists/strobe-cohort/info/#2) | Explain the scientific background and rationale for the investigation being reported | 5 |
| Objectives | [#3](https://www.goodreports.org/reporting-checklists/strobe-cohort/info/#3) | State specific objectives, including any prespecified hypotheses | 5 |
| **Methods** |  |  |  |
| Study design | [#4](https://www.goodreports.org/reporting-checklists/strobe-cohort/info/#4) | Present key elements of study design early in the paper | 5 |
| Setting | [#5](https://www.goodreports.org/reporting-checklists/strobe-cohort/info/#5) | Describe the setting, locations, and relevant dates, including periods of recruitment, exposure, follow-up, and data collection | 5-6 |
| Eligibility criteria | [#6a](https://www.goodreports.org/reporting-checklists/strobe-cohort/info/#6a) | Give the eligibility criteria, and the sources and methods of selection of participants. Describe methods of follow-up. | 5 |
| Eligibility criteria | [#6b](https://www.goodreports.org/reporting-checklists/strobe-cohort/info/#6b) | For matched studies, give matching criteria and number of exposed and unexposed | n/a - not a matched study |
| Variables | [#7](https://www.goodreports.org/reporting-checklists/strobe-cohort/info/#7) | Clearly define all outcomes, exposures, predictors, potential confounders, and effect modifiers. Give diagnostic criteria, if applicable | 6 |
| Data sources / measurement | [#8](https://www.goodreports.org/reporting-checklists/strobe-cohort/info/#8) | For each variable of interest give sources of data and details of methods of assessment (measurement). Describe comparability of assessment methods if there is more than one group. Give information separately for for exposed and unexposed groups if applicable. | 6 |
| Bias | [#9](https://www.goodreports.org/reporting-checklists/strobe-cohort/info/#9) | Describe any efforts to address potential sources of bias | 6 |
| Study size | [#10](https://www.goodreports.org/reporting-checklists/strobe-cohort/info/#10) | Explain how the study size was arrived at | 6 |
| Quantitative variables | [#11](https://www.goodreports.org/reporting-checklists/strobe-cohort/info/#11) | Explain how quantitative variables were handled in the analyses. If applicable, describe which groupings were chosen, and why | 6 |
| Statistical methods | [#12a](https://www.goodreports.org/reporting-checklists/strobe-cohort/info/#12a) | Describe all statistical methods, including those used to control for confounding |  |
| 6-7 |  |  |  |
| Statistical methods | [#12b](https://www.goodreports.org/reporting-checklists/strobe-cohort/info/#12b) | Describe any methods used to examine subgroups and interactions | 6-7 |
| Statistical methods | [#12c](https://www.goodreports.org/reporting-checklists/strobe-cohort/info/#12c) | Explain how missing data were addressed | 6-7 |
| Statistical methods | [#12d](https://www.goodreports.org/reporting-checklists/strobe-cohort/info/#12d) | If applicable, explain how loss to follow-up was addressed | 7 - cases excluded from analysis due to insufficient information |
| Statistical methods | [#12e](https://www.goodreports.org/reporting-checklists/strobe-cohort/info/#12e) | Describe any sensitivity analyses |  |
| 10 |  |  |  |
| **Results** |  |  |  |
| Participants | [#13a](https://www.goodreports.org/reporting-checklists/strobe-cohort/info/#13a) | Report numbers of individuals at each stage of study—eg numbers potentially eligible, examined for eligibility, confirmed eligible, included in the study, completing follow-up, and analysed. Give information separately for for exposed and unexposed groups if applicable. | 7 |
| Participants | [#13b](https://www.goodreports.org/reporting-checklists/strobe-cohort/info/#13b) | Give reasons for non-participation at each stage | 7 |
| Participants | [#13c](https://www.goodreports.org/reporting-checklists/strobe-cohort/info/#13c) | Consider use of a flow diagram |  |
| n/a - only one round of exclusion used in this analysis |  |  |  |
| Descriptive data | [#14a](https://www.goodreports.org/reporting-checklists/strobe-cohort/info/#14a) | Give characteristics of study participants (eg demographic, clinical, social) and information on exposures and potential confounders. Give information separately for exposed and unexposed groups if applicable. | 7 |
| Descriptive data | [#14b](https://www.goodreports.org/reporting-checklists/strobe-cohort/info/#14b) | Indicate number of participants with missing data for each variable of interest |  |
| 16-17 |  |  |  |
| Descriptive data | [#14c](https://www.goodreports.org/reporting-checklists/strobe-cohort/info/#14c) | Summarise follow-up time (eg, average and total amount) |  |
| n/a - follow-up time not calculated |  |  |  |
| Outcome data | [#15](https://www.goodreports.org/reporting-checklists/strobe-cohort/info/#15) | Report numbers of outcome events or summary measures over time. Give information separately for exposed and unexposed groups if applicable. |  |
| 7-8 |  |  |  |
| Main results | [#16a](https://www.goodreports.org/reporting-checklists/strobe-cohort/info/#16a) | Give unadjusted estimates and, if applicable, confounder-adjusted estimates and their precision (eg, 95% confidence interval). Make clear which confounders were adjusted for and why they were included | 7-8 |
| Main results | [#16b](https://www.goodreports.org/reporting-checklists/strobe-cohort/info/#16b) | Report category boundaries when continuous variables were categorized | 16-17 |
| Main results | [#16c](https://www.goodreports.org/reporting-checklists/strobe-cohort/info/#16c) | If relevant, consider translating estimates of relative risk into absolute risk for a meaningful time period |  |
| 16-17 |  |  |  |
| Other analyses | [#17](https://www.goodreports.org/reporting-checklists/strobe-cohort/info/#17) | Report other analyses done—eg analyses of subgroups and interactions, and sensitivity analyses | 10 |
| **Discussion** |  |  |  |
| Key results | [#18](https://www.goodreports.org/reporting-checklists/strobe-cohort/info/#18) | Summarise key results with reference to study objectives | 8 |
| Limitations | [#19](https://www.goodreports.org/reporting-checklists/strobe-cohort/info/#19) | Discuss limitations of the study, taking into account sources of potential bias or imprecision. Discuss both direction and magnitude of any potential bias. | 9-10 |
| Interpretation | [#20](https://www.goodreports.org/reporting-checklists/strobe-cohort/info/#20) | Give a cautious overall interpretation considering objectives, limitations, multiplicity of analyses, results from similar studies, and other relevant evidence. | 8-9 |
| Generalisability | [#21](https://www.goodreports.org/reporting-checklists/strobe-cohort/info/#21) | Discuss the generalisability (external validity) of the study results | 10 |
| **Other Information** |  |  |  |
| Funding | [#22](https://www.goodreports.org/reporting-checklists/strobe-cohort/info/#22) | Give the source of funding and the role of the funders for the present study and, if applicable, for the original study on which the present article is based | 11 |

Notes:

- 6b: n/a - not a matched study
- 12d: 7 - cases excluded from analysis due to insufficient information
- 13c: n/a - only one round of exclusion used in this analysis
- 14c: n/a - follow-up time not calculated The STROBE checklist is distributed under the terms of the Creative Commons Attribution License CC-BY. This checklist was completed on 22. February 2021 using <https://www.goodreports.org/>, a tool made by the [EQUATOR Network](https://www.equator-network.org) in collaboration with [Penelope.ai](https://www.penelope.ai)
